# The timing of contact restrictions and pro-active testing balances the socio-economic impact of a lockdown with the control of infections

**DOI:** 10.1101/2020.05.08.20095596

**Authors:** Saptarshi Bej, Olaf Wolkenhauer

## Abstract

During the SARS-CoV-2 pandemic, numerous mathematical models have been developed. Reporting artefacts and missing data about asymptomatic spreaders, imply considerable margins of uncertainty for model-based predictions. Epidemiological models can however also be used to investigate the consequences of measures to control the pandemic, reflected in changes to parameter values.

We present a SIR-based, SUIR model in which the influence of testing and a reduction of contacts is studied by distinguishing ‘Unidentified’ and ‘Identified’ spreaders of infections. The model uses four ordinary differential equations and is kept deliberately simple to investigate general patterns occurring from testing and contact restrictions. The model goes beyond other efforts, by introducing time dependent parameter curves that represent different strategies in controlling the pandemic.

Our analysis reveals the effect of ‘pro-active’ testing for the design of contact restriction measures. By pro-active testing we mean testing beyond those people who show symptoms. The simulations can explain why the timing of contract restrictions and pro-active testing is important. The model can also be used to study the consequence of different strategies to exit from lockdown.

Our SUIR model is implemented in Python and is made available through a Juypter Notebooks. This an extensive documentation of the derivation and implementation of the model, as well as transparent and reproducible simulation studies. Our model should contribute to a better understanding of the role of testing and contact restrictions.

## 1 Introduction

During the SARS-CoV-2 pandemic, numerous mathematical models have been developed. Using these models to predict infections, for a particular region or country has been very difficult to do a lack of sufficiently rich and time accurate datasets. Reporting artefacts and missing data about asymptomatic spreaders, imply considerable margins of uncertainty for predictions.

There is however a second use of models, that investigate general pattern and can answer questions, including: What is the role of testing in controlling the pandemic? What difference does an early restriction of contacts (lockdown) have on the timing and size of infections have? What effect does the length of the lockdown have on the occurrence of a second peak have?

We present here a [SIR](https://www.nature.com/articles/s41592-020-0822-z)-based, SUIR model motivated by [Singh and Adhikary](https://arxiv.org/pdf/2003.12055.pdf), in which the influence of testing and a reduction of contacts is studied by distinguishing ‘Unidentified’ and ‘Identified’ spreaders of infections [1, 2]. The model uses four ordinary differential equations and is kept deliberately simple to investigate general pattern occurring from testing and contact restrictions. The model goes beyond other efforts, by introducing time dependent parameter curves that represent different strategies in controlling the pandemic.

Our analysis reveals the effect of ‘pro-active’ testing for the design of contact restriction measures. By pro-active testing we mean testing beyond those people who show symptoms. The simulations can explain why the timing of contract restrictions and pro-active testing is important. The model can also be used to study the consequence of different strategies to exit from lockdown, whether a short lockdown, combined with pro-active testing, can be sufficient or whether an exit strategy with a cyclic pattern of restrictions and no restrictions is appropriate. Our simulations also show that a short lockdown or period lockdown-exit pattern bring substantial reductions in the number of infections, without a long-term lockdown of large parts of the economy.

Since both, a total lockdown and pro-active testing will be difficult to achieve, especially in the early phase of a pandemic. Our analysis suggests however that with pro-active testing and strict isolation of identified spreaders, it is possible to achieve control of the pandemic without a total lockdown.

## 2 Compartmental SUIR Modelling

During the initial phase of the SARS-CoV-2 pandemic, most tests conducted were on people showing symptoms. We are here particularly interested in “pro-active” testing, which we interpret to be testing individuals even without symptoms. Pro-active testing may consider the following scenarios:

- Random testing of individuals (for example, walk-in testing stations installed in South Korea)
- Strategic testing such as testing of all persons who have been in contact with some identified patient
- Testing of all persons in identified ‘hot-spots’ of the disease

We assume that the health system is economically and socially capable of organizing an pro-active testing scenario. In such a case, as opposed to the SEIR model, one does not need to consider the the compartment E of exposed individuals, because pro-active testing is done irrespective of the fact whether a person in susceptible, exposed, infected.

Our model is quite similar to the basic [SIR model](https://www.nature.com/articles/s41592-020-0822-z), but with the *I* compartment split into two parts, one capturing identified spreaders, and another un-identified spreaders [1]. The construction of our model can also be considered a simplified version of a model proposed by [Singh and Adhikary](https://arxiv.org/pdf/2003.12055.pdf) [2]. We interpret the model slightly differently to investigate the importance of pro-active testing in controlling the pandemic. We consider the following groups (“compartments”):

- *S*: Susceptible; People who can still become infected
- *U*: Unidentified (silent) spreaders; People who can spread the infection and have not been tested to be COVID positive yet
- *I*: Identified spreaders; People who can spread the infection and have been tested to be COVID positive
- *R*: Resolved; Resolved cases (deaths+recovery)

Note that, we use the notion of Unidentified spreaders and Identified spreaders in this model since the Unidentified spreaders have a large role in spreading the disease. We usually do not know the actual number of infections. During the initial phase of the outbreak, PCR based tests could only detect an active infection. So, we do not consider the Infection compartment as usually done in classical SEIR model. Rather we defined the compartment as ‘Identified spreaders’, a variable that can be reliably evaluated, simply because the primary data usually available publicly is the number of cases appearing each day. Note that, these numbers do not represent the total number of infections or total number of spreaders, but only the number of persons who have been tested. The reported number of identified cases is also highly dependant on the degree of testing performed by a medical system. This model considers a parameter quantifying the probability of a disease spreader to be identified. In a practical scenario, this probability is likely to be high when there is pro-active testing. Moreover, it considers parameters accounting for social isolation assuming that, an identified spreader would be rigorously quarantined and would thus contribute significantly less in spreading the epidemic.

In the next section, we derive the equation of the model step-by-step.

### 2.1 Construction of the model

Let *S*(*t*) count how many individuals are susceptible at time *t*. This group is at risk of becoming infected.

Assuming that most people are not isolated, they can get infected. The initial condition for *S* would thus be approximately *N* − *U*(0) − *I*(0) − *R*(0), where *N* is the size of the population at risk of contracting the virus.

The group *U*(*t*) at time *t* are unidentified (silent) spreaders - people who are contagious, who can spread the infection but have not been tested to be positive yet.

Any increase to *U*, implies an equal decrease of *S*. Any term that adds to the equation for *U*, will be mirrored by the same term with a minus in front in the equation for *S*.

The group of identified spreaders at time *t* is denoted *I*(*t*). The individuals have been tested positive, and can spread the disease. Once tested positive, one can assume that these person will be isolated reasonably well. This means the contribution of this group to the decrease of *S* should be small, compared to the contribution from *U*. We shall denote these contributions of *U* and *I* to the decrease of *S* with *κ_I_* and *κ_U_*

We refer to *κ_I_* as the **Isolation Index**, defining the effectiveness of isolating identified spreaders. A low value of *κ_I_* refers to good quarantining, through strict isolation of identified spreaders.

The **Contact restriction Index** *κ_U_* defines the contribution of unidentified spreaders on the infection of susceptibles. If measures, like contact restrictions are implemented, their effectiveness would be reflected by *κ_U_*. A low value of *κ_U_*, refers to strict contact restrictions as would happen during a lockdown.

One would then expect that *κ_U_* ≫ κ*_I_*

Any increase of identified spreaders *I*, implies an equal decrease of *S*. Any term that adds to the equation for *I*, will be mirrored by the same term with a minus in front in the equation for *S*.

Finally, we combine death and recoveries in the subpopulation denoted by *R*(*t*). Considering subpopulations *S*, *U*, *I*, and *R*, we may refer to our model as a SUIR model. Models, where individuals move from one subpopulation “compartment”) to another, are also referred to as ‘compartmental models’. Most classical epidemiological models include *E* for an exposed group, which means that our model slightly deviates from standard models. We deliberately avoided a larger model with more variables. The current coronavirus pandemic has revealed various problems with data generated. The larger the size of the model, the greater the risk of making it unidentifiable. The predictions arising from SEIR type epidemiological models also tend to be sensitive to changes in parameter values. Taken together, during the current crisis numerical predictions of case numbers and specific times (e.g. of peaks) has been difficult. As a consequence, we here focus on a simple model that does not try to focus on values and time points. Instead, the goal is to investigate general pattern and scenarios. An example of such type of modelling that we pursue here is the model by Uri Alon [3].

### 2.2 Building the equations

Denoting by *t* time in days, the model that looks at daily changes, where Δ*t*=1, would be represented by difference equations. In epidemiological modelling it is however common to assume Δ*t* to go to zero, so that the model is formulated in terms of differential equations. To then simulate the differential equation model, one introduces a uniform mesh in the, *t_n_* = *n*Δ*t*, *n* = 0,..., *N_t_*, and seek *S* at the mesh points. The numerical approximation of *S* at time *t_n_* is denoted by *S^n^*.

In the time interval Δ*t* some people will be infected, so *S* will decrease. Both, *I* and *U* contribute to this decrease,

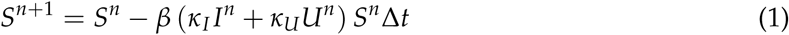

From what we said above, the equation for *I^n^*^+1^ and *U^n^*^+1^ will automatically receive the term,

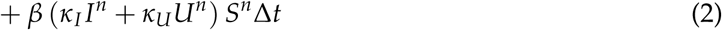

The parameter *β* reflects how easily people get infected during a time interval of unit length (here per day). This is also referred to as the “transmission rate”.

Suppose that during a time interval *T* we measure that *x* actual pairwise meetings do occur among *y* theoretically possible pairings of people from the *S*, *U* and *I* subpopulations. The probability that people meet in pairs during time *T* is, estimated as a relative frequency, *y*/*x*. From such a statistic we want the probability per unit time, *µ* = *x*/(*yT*).

Given the probability *µ*, the expected number of meetings per time interval of (*κ_I_I^n^* + *κ_U_U^n^*)*S^n^* possible pairs of people is *µ*(*κ_I_I^n^* + *κ_U_U^n^*)*S^n^*. During a time interval Δ*t*, there will be *µ*(*κ_I_I^n^* + *κ_U_U^n^*)*S^n^*Δ*t* expected number of meetings between susceptibeles, with identified and unidentified people.

Only a fraction of the *µ*(*κ_I_I^n^* + *κ_U_U^n^*)*S^n^*Δ*t* meetings are effective in transmitting the virus. Counting that *m* people get infected in *n* such pairwise encounters (e.g. 5 are infected in 1000 encounters), we can estimate the probability of being infected as *p* = *x*/*y*. The expected number of individuals in the *S* subpopulation of susceptibles that in time interval Δ*t* catch the virus and get infected is then,

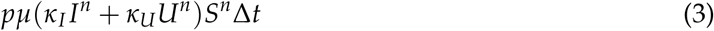

Next, describing the evolution of *I*(*t*), we have from the equation of *S* already one term,

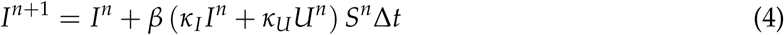

However, there will also be people that either recover, or die, giving us,

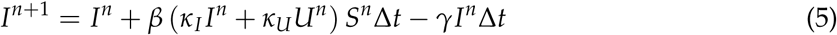

Introducing testing, there will be a proportion *ρ* and *∊* of individuals identified by ‘proactive testing’ of people without symptoms and testing due to the manifestation of symptoms, respectively. This proportion will move to subpopulation *I*. The rest of them moves to compartment *U*. This gives us for *I^n^*^+1^,

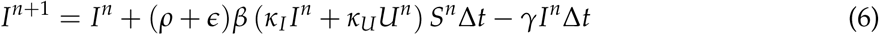

The equation for *U^n^*^+1^, can now be constructed from the symmetry that must be there from the other equations,

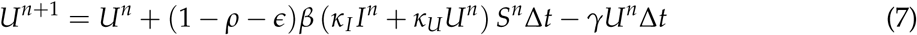

simply be determined by,

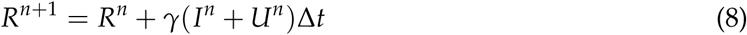

To summarise, we the model is constructed from the following difference equations:

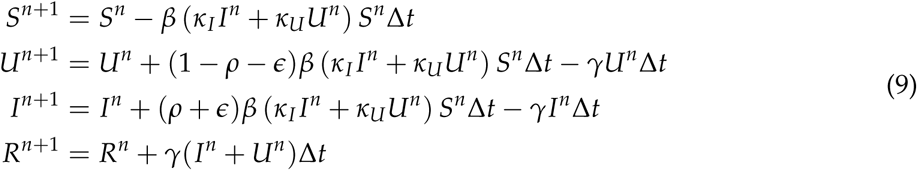

Subtracting the *S^n^*, *U^n^*, *I^n^*, *R^n^* on both sides, and dividing by Δ*t*, gives us the discretitzed version of differential equations, using the Forward Euler method for *n*=0, …, *N_t_*, over some finite interval [0, *T*]. The corresponding ordinary differential equations are:

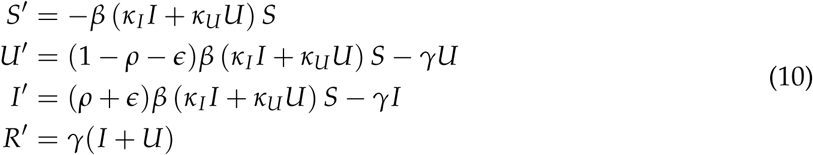

#### Estimating parameter values

For the purpose of our study we shall adopt for initial conditions and values chosen for the key parameters, from the [Nature publication](https://www.nature.com/articles/s41592-020-0822-z) on SIR modelling from 20th April 2020.

The parameter *β* reflects how easily people get infected during a time interval of unit length. This is also referred to as the “transmission rate”. *β*^−1^ is the typical/average time between contacts [Bjoernstad et al Nature April](https://www.nature.com/articles/s41592-020-0822-z) 2020[1].

We refer to *γ* as the **Recovery Index**. It is estimated as the inverse of the average duration of illness. The average duration of illness is assumed to be 14 days [Bjoernstad et al Nature April](https://www.nature.com/articles/s41592-020-0822-z) 2020 [1]. For the SIR model, the relationship between *β* and *γ* is the reproduction number 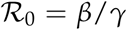. The basic reproduction number *R*(0) represents how many new infections are caused at the start of an epidemic by a spreader. Note that this estimation of 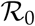 also holds for our SUIR model (See Appendix 5.1). The reason behind is that, the sum of the compartments *U* and *I* essentially is essentially same as the compartment *I_SIR_* representing spreaders in the SIR model. The chance of transmission is directly proportional to the contact frequency, i.e., how many close contacts a person makes on an average per day. During the COVID-19 pandemic, the reproduction number has been adopted in several countries as a criteria for lifting contact restrictions measures. For the estimation of 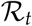 several methods exist and depending on how it is calculated, the estimates can differ quite considerably. In Germany, the Robert Koch Institute has estimated 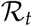 as the quotient of new daily infections in two successive four day windows. The model from Michael Meyer-Herrmann from the Helmholtz Centre, Braunschweig, also used a sliding window, of seven days, but also included a [model](https://gitlab.com/simm/covid19/secir/-/wikis/Report) in the estimate of 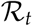 [4].

A virus incubates for some time after it enters a human but before it causes symptoms. The incubation period for the new coronavirus SARS-CoV-2 typically ranges from two to 14 days, with the median being four or five days. During some of the incubation period, a person can be infectious. The parameter *€* can be interpreted as the **inverse of the incubation period** (in days). We assume this to be around 5 days [5]. The incubation period is the average number of days after which a person shows perceivable symptoms and is therefore forced to take a COVID test. *∊* is thus linked to testing focusing on those with symptoms, and a low *∊*, corresponds to a longer incubation period, which means that more spreaders will be unidentified.

**Pro-active Testing Index** *ρ* describes the proportion of spreaders identified as COVID +ve. This parameter relates to the aggressiveness of testing. We assume that the more pro-active the testing is, the higher the probability of the detection of a previously unidentified spreader. A low value of *ρ*, corresponds to little aggressive testing, implying more unidentified spreaders. A high value of *ρ*, describes pro-active testing, reducing the number of unidentified spreaders.

Finally, the population size *N* is often chosen for the initial condition of the susceptibles group *S*(0). But even this figure can be debated. During the SARS-CoV-2 pandemic, the spread across countries occurred through people travelling and then within countries primarily linked to more densely populated regions. I would therefore not make much sense to choose the population size of a country to decide upon the initial condition for *S*.

Initial conditions and other assumptions:

We assume that in the beginning of the pandemic, in a hot-spot, there are 20 unidentified spreaders per population of 100,000 and 1 identified spreader per million. Visualizing the results, we display counts per 1 million. The reason is the population of a standard administrative unit (a country or state) is of the same order. We simulate our model for a period of 365 days considering each time unit, a day. Since our focus is testing and contract restrictions, linked to identified and unidentified spreaders, we shall hereafter only plot *I* and *U*, or the sum of spreaders, *U* + *I*. We emphasize again here that our model is deliberately not trying to make accurate predictions about cases numbers at a particular point in time, for a specific region or country. We are thus not fitting our model to actual data for a particular region. The focus is on studying general patterns, arising from different scenarios, combining contact restrictions with testing.

## 3 Results and discussions

In this Section present our results with follow up discussions in Subsections 3.1, 3.2 and 3.3. In Subsection 3.1, we investigate different scenarios of contract restrictions and pro-active testing. In these scenarios parameters are not time-dependant:

- Scenario 1: No isolation of identified spreaders; No contact restrictions; No pro-active testing
- Scenario 2: Isolation of identified spreaders; No contact restrictions; No pro-active testing
- Scenario 3: Isolation of identified spreaders; Contact restrictions; No pro-active testing
- Scenario 4: Isolation of identified spreaders; Contact restrictions; Pro-active testing

In Subsection 3.2, we then introduce time-dependent changes to testing and contact reduction measures, focusing on the timing of these:

- Policy 1: Early lockdown; No pro-active testing
- Policy 2: Late lockdown; No pro-active testing
- Policy 3: No lockdown; Early pro-active testing
- Policy 4: Late lockdown; Late pro-active testing
- Policy 5: Early lockdown; Early pro-active testing
- Ploicy 6: Early lockdown; Late pro-active testing

Finally in Subection 3.3, we compare the following exit strategies from lockdown:

- Strategy 1: Early abrupt exit, Minimal pro-active testing
- Strategy 2: Early gradual exit, Minimal pro-active testing
- Strategy 3: Late abrupt exit, Minimal pro-active testing
- Strategy 4: Late gradual exit, Minimal pro-active testing
- Strategy 5: Late gradual exit, Early pro-active testing with exit
- Strategy 6: Periodic lockdown, Early pro-active testing without exit
- Strategy 7: Periodic lockdown, Late pro-active testing without exit

### 3.1 Scenarios w.r.t isolation, contact restrictions, and pro-active testing

For the following simulation studies we consider different scenarios with changes to *κ_I_* (isolation), *κ_U_* (contact restrictions), and *ρ* (pro-active testing). The parameters *γ*, *β* and *∊* remain fixed (as are the initial conditions and population size). For the following analyses, the transmission rate *β*, the recovery index *γ*, the inverse of the incubation period *∊*, and the isolation index *κ_I_* will be “fixed”, while the focus of our attention is on the contact restriction index *κ_I_* and pro-active testing index *ρ*. A low value of *κ_U_*, refers to strict contact restrictions as would happen during a lockdown. A high value of *ρ*, describes pro-active testing, reducing the number of unidentified spreaders. Both, *ρ* and *κ_U_* take values in the unit interval [0, 1].

#### Scenario 1: No isolation of identified spreaders; No contact restrictions; No pro-active testing

This scenario corresponds to the situation where there is no isolation of identified spreaders which does not usually happen in practice. This translates to *κ_U_* = 1, *κ_I_* = 1 and *ρ* = 0 in terms of parameter values. Thus, although unrealistic, this is the worst-case scenario can theoretically happen. Here we see the peak to be appearing at around day 120 with more than 150K (120K+30K) spreaders per million population. (See Figure 1)

**Figure 1:**
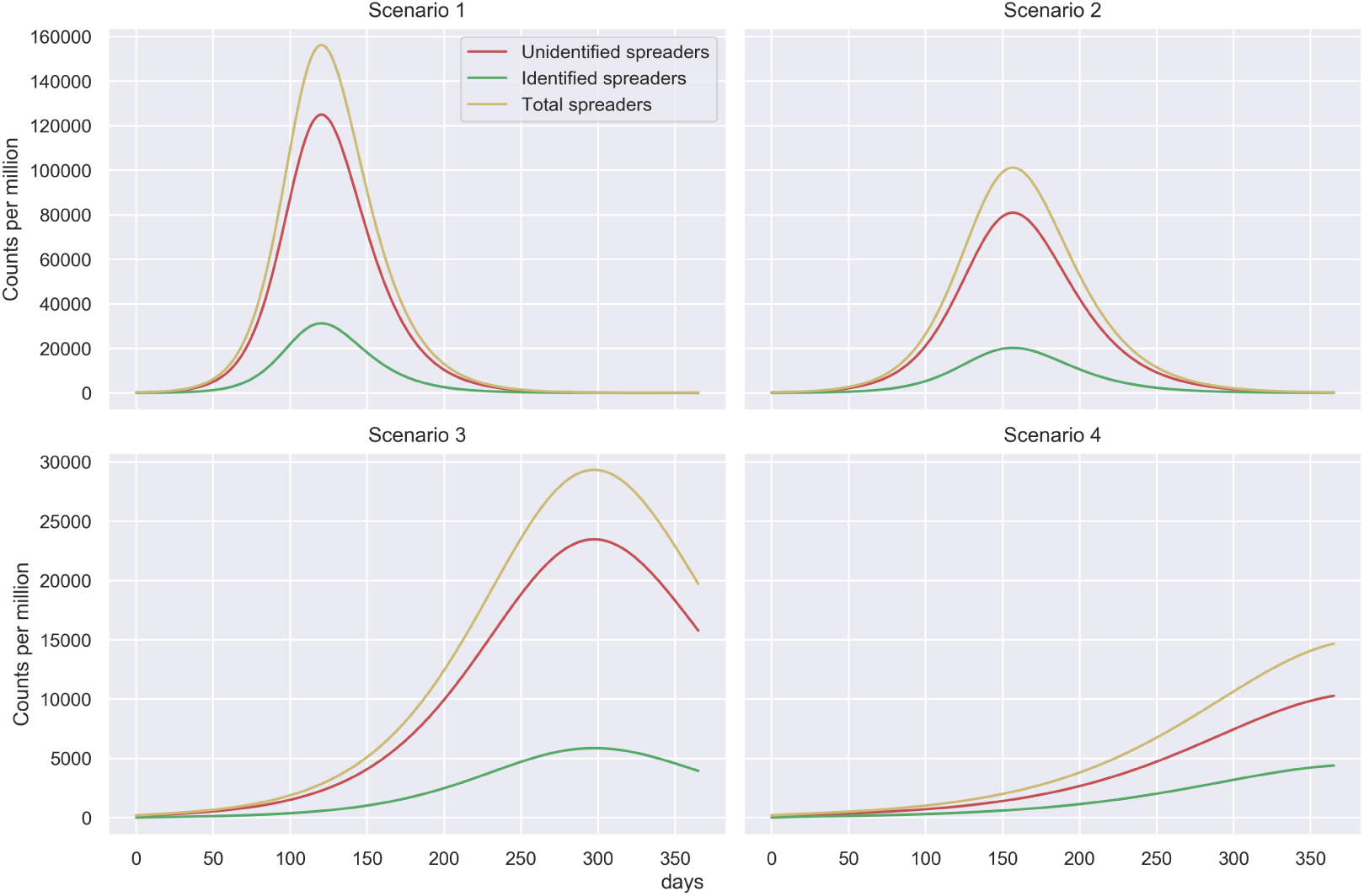
Figure showing number of spreaders for several scenarios

#### Scenario 2: Isolation of identified spreaders; No contact restrictions; No pro-active testing

This scenario corresponds to the situation where there is isolation of spreaders, no contact restriction and no pro-active testing. This translates to *κ_U_* = 1, *κ_I_* = .25 and *ρ* = 0 in terms of parameter values. This scenario is more realistic compared to Scenario 1. Usually any patient with a contagious disease is quarantined by common practice. However, this scenario implies no additional measures such as contact reduction or pro-active testing that can be employed to control a pandemic. We notice here that the peak appears on day 160 with about 100K(80K+20K) spreaders per million population. (See Figure 1)

#### Scenario 3: Isolation of identified spreaders; Contact restrictions; No pro-active testing

This scenario corresponds to the situation where there is isolation of spreaders, contact restriction and no pro-active testing. This translates to *κ_U_* = .75, *κ_I_* = .25 and *ρ* = 0 in terms of parameter values. The assumption of contact reduction of only 25% is however time independent. This might reflect the situation in a county or state with relatively less human contacts that can be attributed to population density or lifestyle. Time independent contact rate thus makes this more of a hypothetical scenario. In this case the peak appears much later around day 300 with about 25K(20K+5K) spreaders per million. This reflects that a population with intrinsically less contact rate is likely to be at a lesser risk. (See Figure 1).

#### Scenario 4: Isolation of identified spreaders; Contact restrictions; Pro-active testing

This scenario corresponds to the situation where there is isolation of spreaders, contact restriction and pro-active testing. This translates to *κ_U_* = 1, *κ_I_* = .25 and *ρ* = .1 in terms of parameter values. The assumption of contact reduction and pro-active testing are however time independent which again makes this a hypothetical scenario. In this case, the peak appears even later compared to Scenario 3 after 1 year timescale. This in general shows that a 10% pro-active testing (detection of 10% of the unidentified spreaders) can have a considerable effect in further ‘flattening the curve’. It is worthwhile to mention here, that this is an important observation given that the pandemic re-surges after some time. If we can come up with a stable and affordable testing method before the second resurgence, we would be at a much lower risk. (See Figure 1).

#### Comparing Scenarios

What we see in the Figure 2, showing the total number of spreaders (*U*+*I*), for the different scenarios, is the following. Scenario 1, is the worst case, with little action taken to respond to the pandemic. Scenario 2 gives people maximum freedom of movement following by strict isolation of infected individuals. The peak appears a little later compared to Scenario 1. Scenario 3, implements measures for contact restrictions, with good isolation of identified spreaders but no pro-active testing. Scenario 4, in addition to conditions of Scenario 3 there is some testing beyond people with symptoms (pro-active testing).

**Figure 2:**
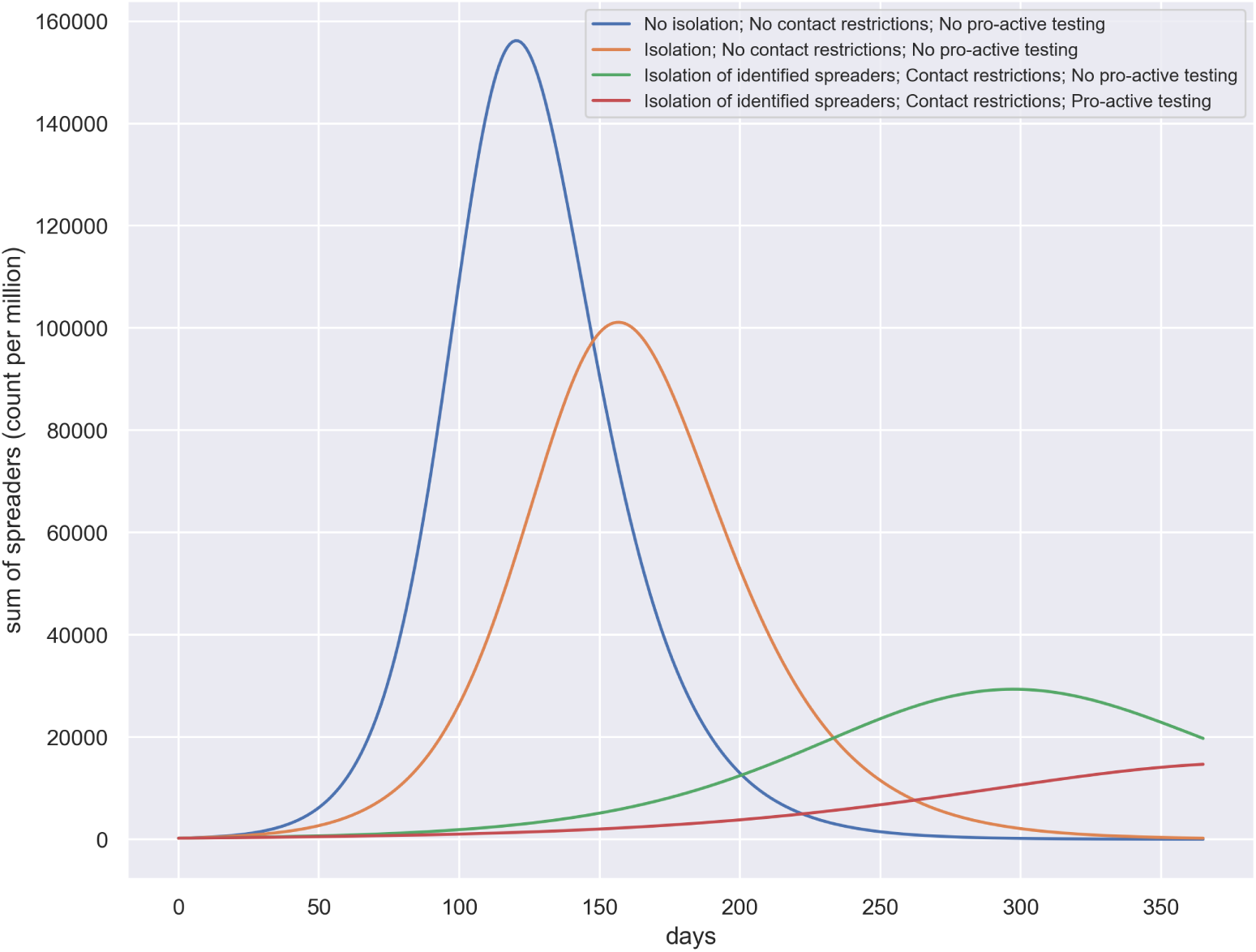
Figure showing for each scenario the temporal evolution of all spreaders, ie the sum of identified and unidentified spreaders.

Our analysis shows that Scenario 4 reflects best management of the pandemic in terms of ‘flattening the curve’. The simulation confirms what other simulation studies have already shown numerous times - contract restrictions help flattening the curve and shifting the peak.

However, the simulation study above, like most others, assume static values for parameters *κ_I_*, *κ_U_* and *ρ*. This might not reflect the dynamics of the pandemic, including the implementation of policies and response of the population to the pandemic. For example, if we assume *κ_U_* = 0.75, we assume that from the beginning itself there is a restriction on social interactions imposed, which is quite far from reality. For this reason, we next observe the dynamics of our pandemic model for time dependent parameters *κ_U_* and *ρ*. We do not assume time dependent *κ_I_* since, for any contagious disease (even normal flu) a diagnosed patient is quarantined anyway. We translate several time dependent parameter settings to real-world scenarios.

### 3.2 Analysing late/early implementation of policies

We here consider time dependent changes in testing and contact restrictions. We introduce curves derived from the logistic function to model time dependent changes in *κ_U_* (linked to contact restrictions) and *ρ* (linked to pro-active testing). Other parameters are not changed to compare the policies: *κ_I_* = 0.2, *β* = 0.14, *γ* = 0.071, *∊* = 0.2.

#### Modeling with time dependent parameters

We create two functions, one modelling an increasing parameter value and another a decreasing parameter value.

For example, most countries had initially free social interactions with no restrictions, which corresponds to a higher *κ_U_* at the beginning of the pandemic. As the awareness about the pandemic increases and with lockdown measures implemented, social interactions are limited. We model this scenario with a flipped logistic function.

Similarly, for most countries testing would pick up over time. pro-active testing, which goes beyond testing people with symptoms, will start delayed. We model this by a time dependent *ρ* value, following the shape of a logistic function.

To compare policies in different countries, we can shift the inflection point forward or backward. For example, comparing the UK with Germany, it is widely acknowledged that widespread testing started earlier in Germany. For social distancing, comparing Norway and Sweden, Norway started social distancing much earlier than Sweden. With these policy functions above we can cover a wide range of policies through which a country or region might respond to the pandemic.

#### Assumptions on parameters for the implementation of policies

The COVID-10 pandemic has effected numerous countries. After about two months, the effects of different policies to deal with the pandemic at national level have emerged. Comparing the UK with Germany, it is widely acknowledged that Germany responded faster with testing and contact restriction measures. Comparing Sweden to Norway, there was a notable difference to when social distancing was enforced. (While Sweden did not enforce contact restriction measures, when the number of COVID-10 related death were rising, the behavior of people changed). Two primary aspects of a pandemic related policy, are

- promptness in implementing a lockdown (total or partial)
- promptness in ramping up the testing procedure

There are other policies that an administrative system could adopt, such as ramping up hospital beds, and ICUs, but these are more related to dealing with the effect of the pandemic while not so much to controlling the spread of the pandemic.

To describe the policies as we implement in our model in terms of numbers, the following points should be noted:

- By early lockdown, we mean that the lockdown is enforced at a timeline of approximately 40 days with contact restriction index of 0.3.
- By late lockdown, we mean that the lockdown is enforced at a timeline of approximately 60 days with contact restriction index of 0.3.
- By early pro-active testing, we mean that the pro-active testing starts at around 60 days with pro-active testing index of 0.7.
- By late pro-active testing, we mean that the pro-active testing starts at around 90 days with pro-active testing index of 0.7.

#### Policy 1: Early lockdown; No pro-active testing

This policy is the most realistic and have been adopted by many countries/states as most counties/states do not have the possibility to have a high rate of pro-active testing. We note here that the peak appears at around 45 days with around 1300 spreaders per million. (See Figure 3)

**Figure 3:**
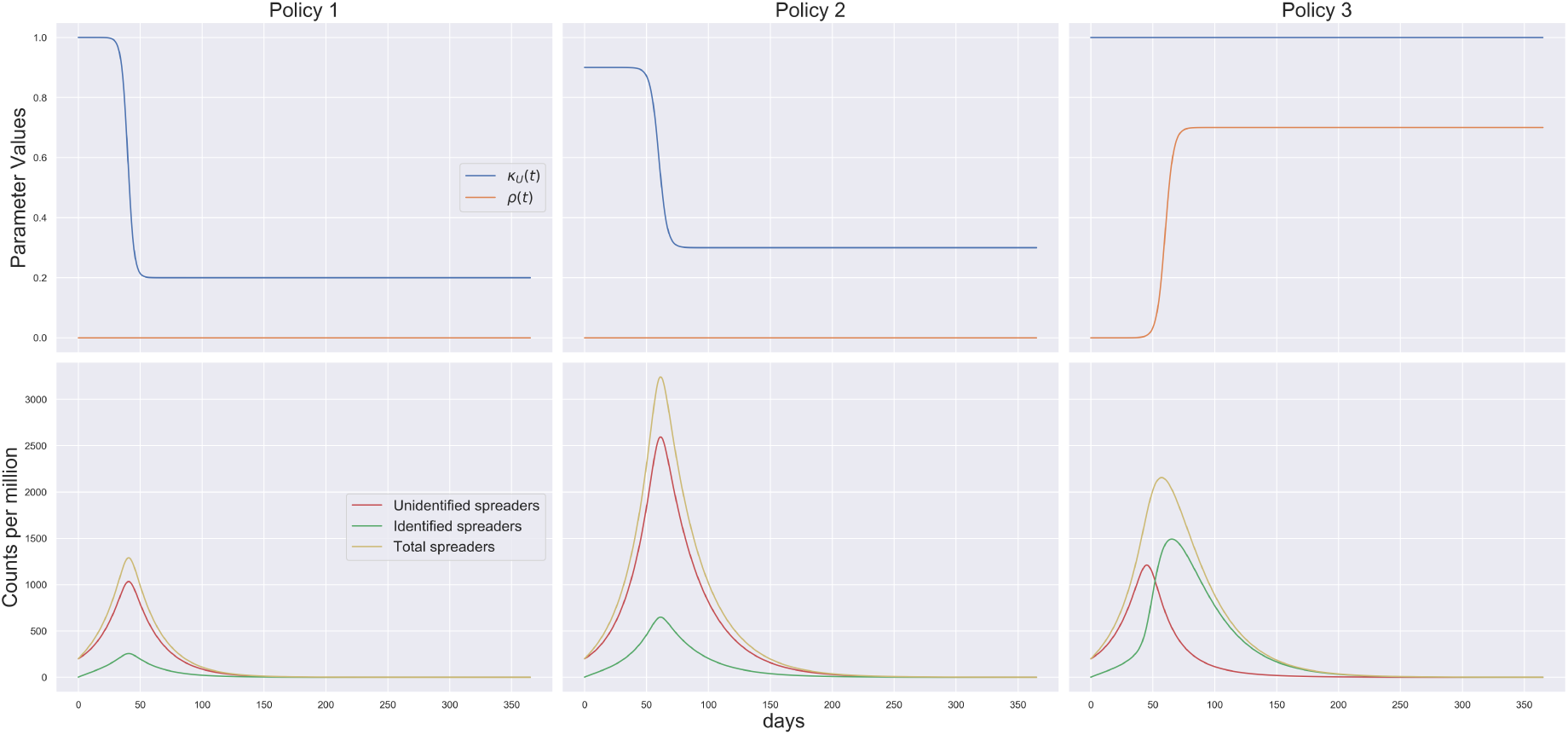
Figure showing parameter values (first row) and number of spreaders (second row) for Policies 1-3 (represented column-wise)

#### Policy 2: Late lockdown; No pro-active testing

This policy reflects the possible situation in some countries/states who, might not have been prompt enough with their lockdown policy. By late lockdown, we mean that the lockdown is enforced at a timeline of approximately 60 days which is 20 days later compared to early lockdown. We note here that the peak appears at around 65 days with around more than 3000 spreaders per million. Comparing to Policy 1, we can thus conclude that a lockdown delay of 20 days might increase the number of spreaders more than twofold. (See Figure 3)

#### Policy 3: No lockdown; Early pro-active testing

This policy at this point is hypothetical. However if there is future relapse of the pandemic at a time when a pro-active testing is affordable, this scenario can be of importance. We note here that the number of identified spreaders becomes higher than the number of unidentified spreaders at around 50 days. The peaks for the identified and unidentified spreaders occur at around 65 days and 45 days with 1500 and 1200 spreaders respectively. The peak number of total spreaders is around 2200 on around 55 days. This shows that if pro-active testing can be employed, even without any lockdown, the effect of the pandemic can be considerably diminished. (See Figure 3)

#### Policy 4: Late lockdown; Late pro-active testing

For this policy the peak appears at around 55-60 days with around 3000 (2500+500) spreaders. The delay in lockdown clearly could not be compensated with the pro active testing (which also starts late). Interestingly, this policy has more number of spreaders at its peak compared to Policy 3 where there was no lockdown at all and early pro-active testing. (See Figure 4)

**Figure 4:**
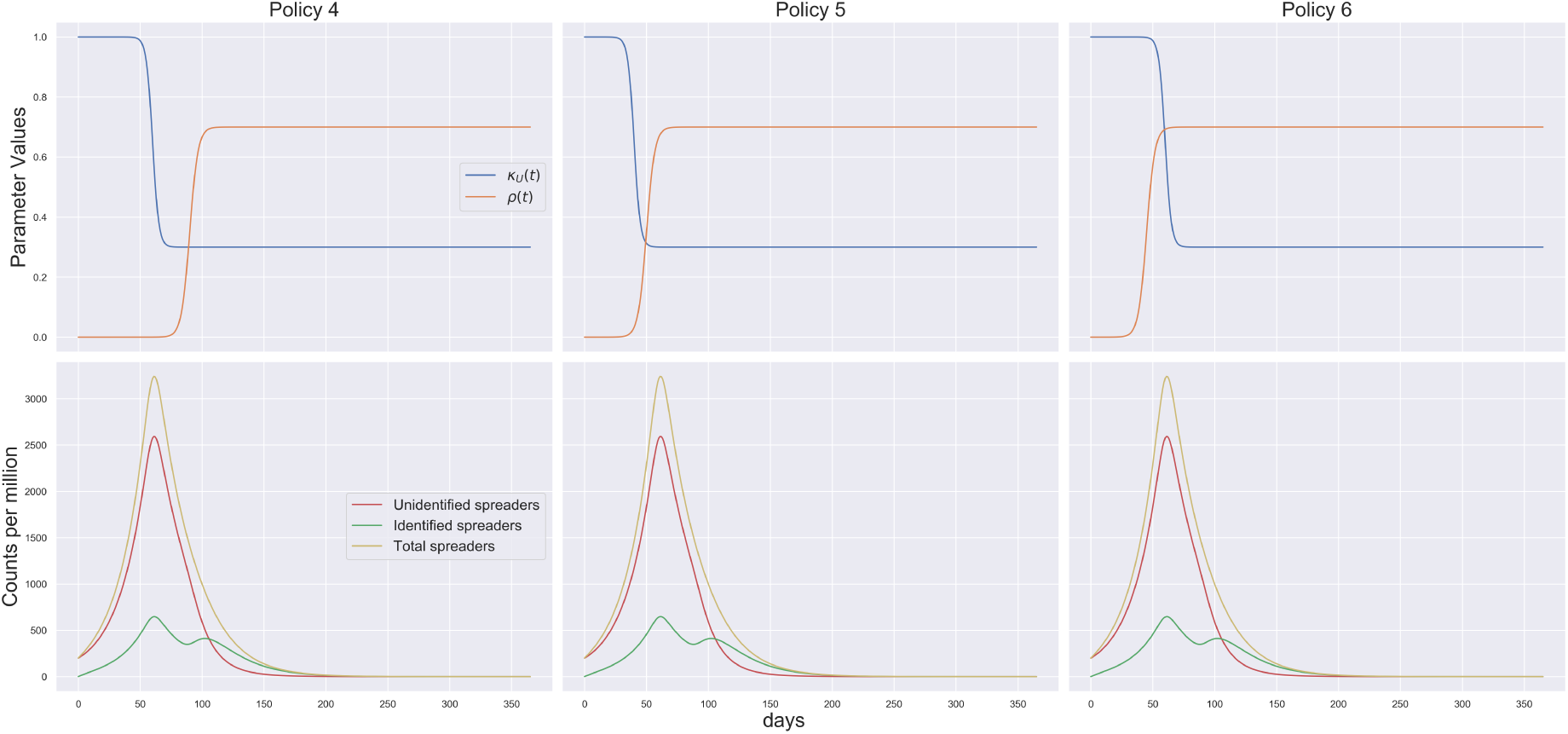
Figure showing parameter values (first row) and number of spreaders (second row) for Policies 4-6 (represented column-wise)

#### Policy 5: Early lockdown; Early pro-active testing

This policy has been employed by South Korea to some extent. The peaks for the identified and unidentified spreaders occur at around 55 days and 45 days with 400 and 1000 spreaders respectively. Note that number of identified spreaders peak, around the time when the testing starts. Thus the peak of the identified spreaders shift to right compared to Policy 1, which corresponds to early lockdown and no proactive testing. Considering the total number of spreaders, this would be the most successful entry policy to the pandemic.(See Figure 4)

#### Policy 6: Early lockdown; Late pro-active testing

This scenario is also likely to happen in practice, given that we one develops a pro-active testing scenario relatively later, which is more likely. The peak for this policy is around day 45 with around 1300 (1050+250) spreaders. Note that number of identified spreaders show a smaller peak, around the time when the testing starts. Considering the total number of spreaders, this would be a fairly successful entry policy to the pandemic. (See Figure 4)

#### Policy 7: Late lockdown; Early pro-active testing

The situation described by this policy is hypothetical and very unlikely to happen. The peak for this policy is around day 45 with around 1300 (1050+250) spreaders.

#### Comparing several entry policies for the pandemic

From the comparative plot of the policies in Figure 5, we notice that there is one group of policies leading to the same peak at aound 40 days and less than 1500 spreaders per million. This includes the following policies:

- Early lockdown; No pro-active testing
- Early lockdown; Early pro-active testing
- Early lockdown; Late pro-active testing
- Late lockdown; Early pro-active testing

We can thus argue that a late lockdown can only be compensated for with early pro active testing. Early on in a pandemic it is however unrealistic to have available large scale random testing. For these simulation studies we consider lockdown strategies that remain implemented. In the next section we shall investigate the question when it is best to relax a lockdown.

### 3.3 Analysing exit strategies from lockdown

As we noticed above, ‘Early lockdown and no Pro-active testing’ strategy is the one that was implemented during the pandemic by numerous countries, from around the middle of March 2020. We shall therefore consider the corresponding peak as a reference and starting point to discuss the timing of relaxation measures/exit strategies. A major motivation behind studying exit strategies is to observer whether there is a chance of relapsing of the pandemic depending on exit strategies. Note that, we have assumed that for each of the exit strategies the entry policy to the pandemic is an early lockdown (at 40 days). There are several variations of contact reduction scenarios that we compare:

- Early abrupt exit
- Early gradual exit
- Late abrupt exit
- Late gradual exit
- Periodic exit

**Figure 5:**
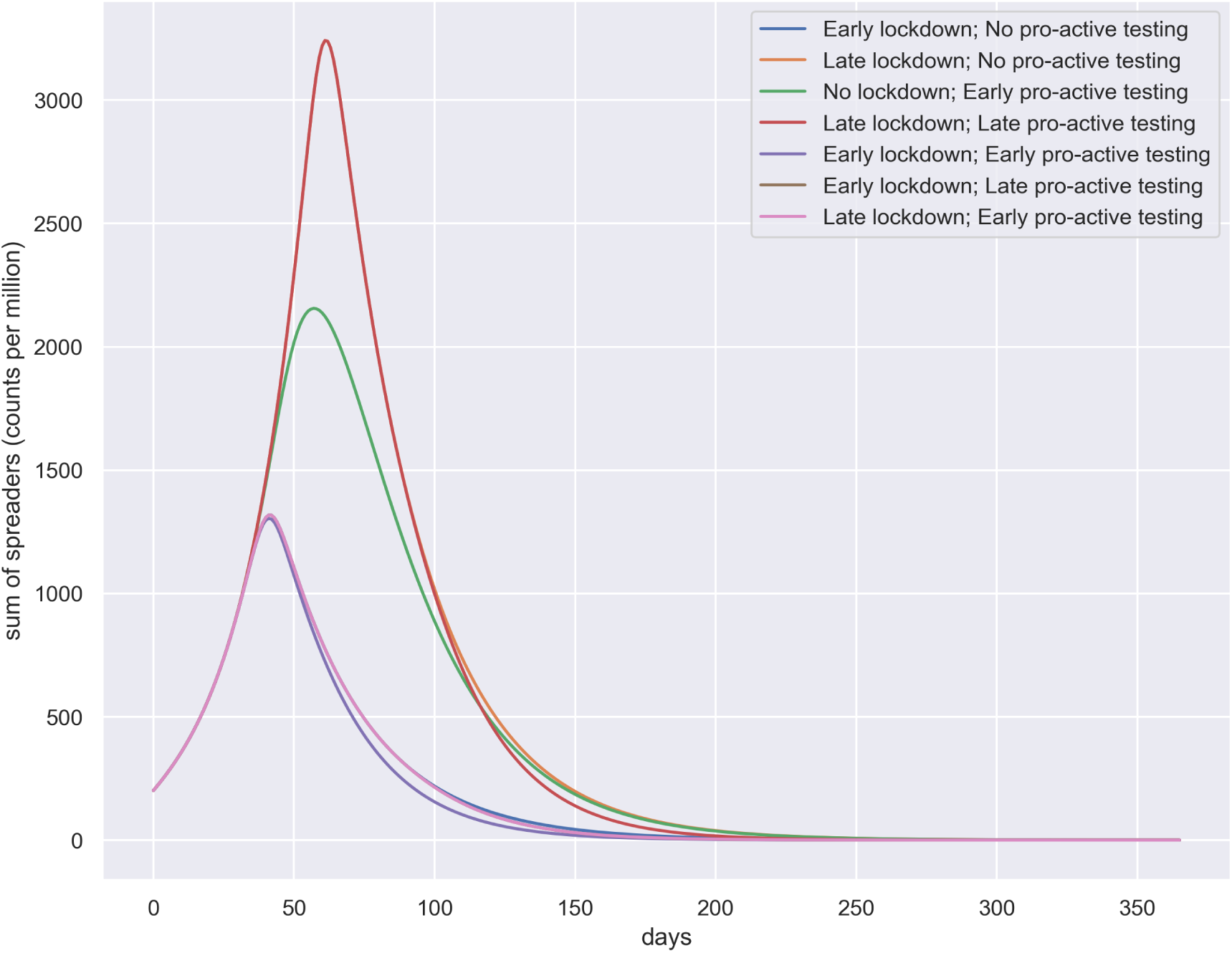
Figure showing for each policy the temporal evolution of spreaders, ie the sum of identified and unidentified spreaders for several policies.

Moreover, there are two variations of pro-active testing scenarios we compare:

- Minimal pro-active testing
- High pro-active testing

We build each strategy as a combination of these variations of contact reduction scenarios and pro-active testing scenarios.

#### Strategy 1: Early abrupt exit; Minimal pro-active testing

Early abrupt exit; Minimal pro-active testing: This corresponds to exiting the lockdown between days 82-92. It is a sudden lift of the lockdown after a lockdown period of ~ 6 weeks. The normalcy returns after a time span of about 1 week. There is a minimal amount of pro active testing considered, which is usually closer to reality. This shows that after achieving a first peak at around 40-45th day, the pandemic subsides. But it again starts relapsing from around 120th day. Since we assume that at the time of second relapse there is no contact reduction whatsoever, the situations turns out to be quite similar to Scenario 2 discussed before. The peak number 80K (60K+20K) appearing at around day 265 is a bit less than that of Scenario 2 because of the minimal pro-active testing happening in this case as opposed to Scenario 2. (See Figure 6)

**Figure 6:**
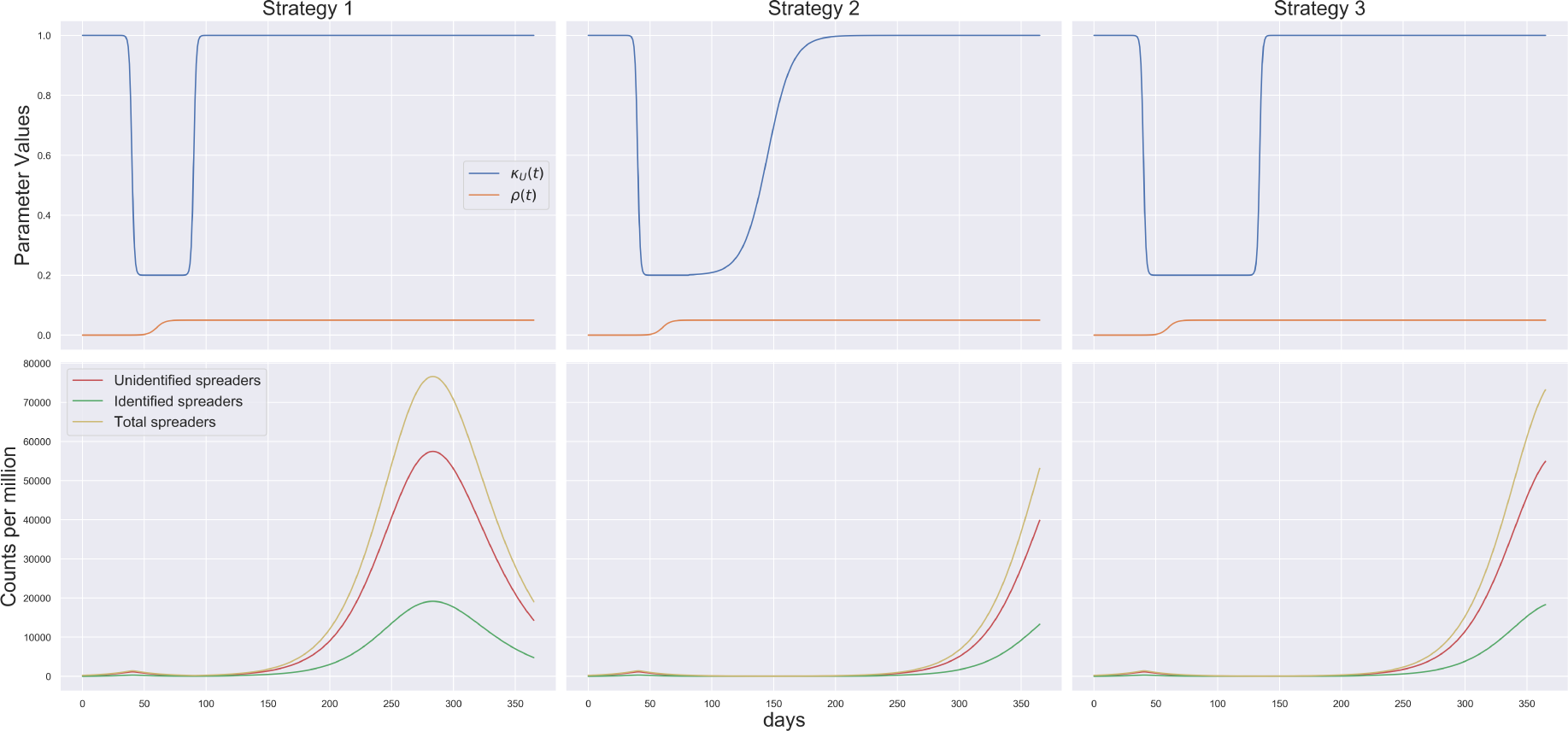
Figure showing parameter values (first row) and number of spreaders (second row) for Strategies 1-3 (represented column-wise)

#### Strategy 2: Early gradual exit; Minimal pro-active testing

Early gradual exit; Minimal pro-active testing: This corresponds to exiting the lockdown between 82-200 days span. It is a gradual lift of the lockdown after a lockdown period of ~ 12 weeks. The normalcy returns after a time span of around 4 months. This shows that after achieving a first peak at around 40-45th day similar to Strategy 1, the pandemic subsides. But it again starts relapsing from around 220th day, much later compared to scenario one, because of the gradual lockdown exit. Except for the starting point and relative transformation of the peak, the relapse peak appearing is exactly the same in structure as of Strategy 1. (See Figure 6)

#### Strategy 3: Late abrupt exit; Minimal pro-active testing

Late abrupt exit; Minimal pro-active testing: This corresponds to exiting the lockdown between days 124-134. It is a sudden lift of the lockdown after a lockdown period of ~ 12 weeks. The normalcy returns after a time span of about 1 week. The situation here is also similar that of Strategy 1, 2. (See Figure 6)

#### Strategy 4: Late gradual exit; Minimal pro-active testing

Late gradual exit; Minimal pro-active testing: This corresponds to exiting the lockdown between days 124-240. It is a gradual lift of the lockdown after a lockdown period of ~ 12 weeks. The normalcy returns after a time span of about 4 months. The situation here is also similar that of Strategy 1, 2 and 3 except for the fact that the relapse of the pandemic occur later in time due to implementation and gradual withdrawal. We note here that even exercising a double lockdown period to that of Strategy 1 and then a gradual exit also might not be enough to prevent future relapses, if we assume that the virus retains the same spreading potential and there is a low degree of pro-active testing. We thus conclude that there is a chance that the pandemic might relapse, within the year after lifting of a short or long lockdown, whether the lockdown is lifted abruptly or gradually, if we assume that the virus retains the same spreading potential and pro-active testing is not boosted up. (See Figure 9)

#### Strategy 5: Late gradual exit; High pro-active testing

From Scenarios 1-4 we noticed that the pandemic might relapse within a year irrespective of when or how the lockdown is lifted assuming that the spreading potential of the virus is maintained and there is little pro-active testing. In this case we see that even if there is short lockdown and an abrupt exit, a high pro-active testing index of .7 can prevent the pandemic from relapsing. Recall here that pro-active testing sho. Recall here that pro-active testing should not be confused with random testing. That is, *ρ* = 0.7 does not mean randomly testing 70 percent of the population. Rather it means implementation of testing strategies that can ensure that 70 percent of the spreaders are detected. This can be done by:

* Rigorous testing of persons in close contact with an identified spreader * Random testing in identified hotspots

(See Figure 7) This can be done by:

- Rigorous testing of persons in close contact with an identified spreader
- Random testing in identified hotspots

**Figure 7:**
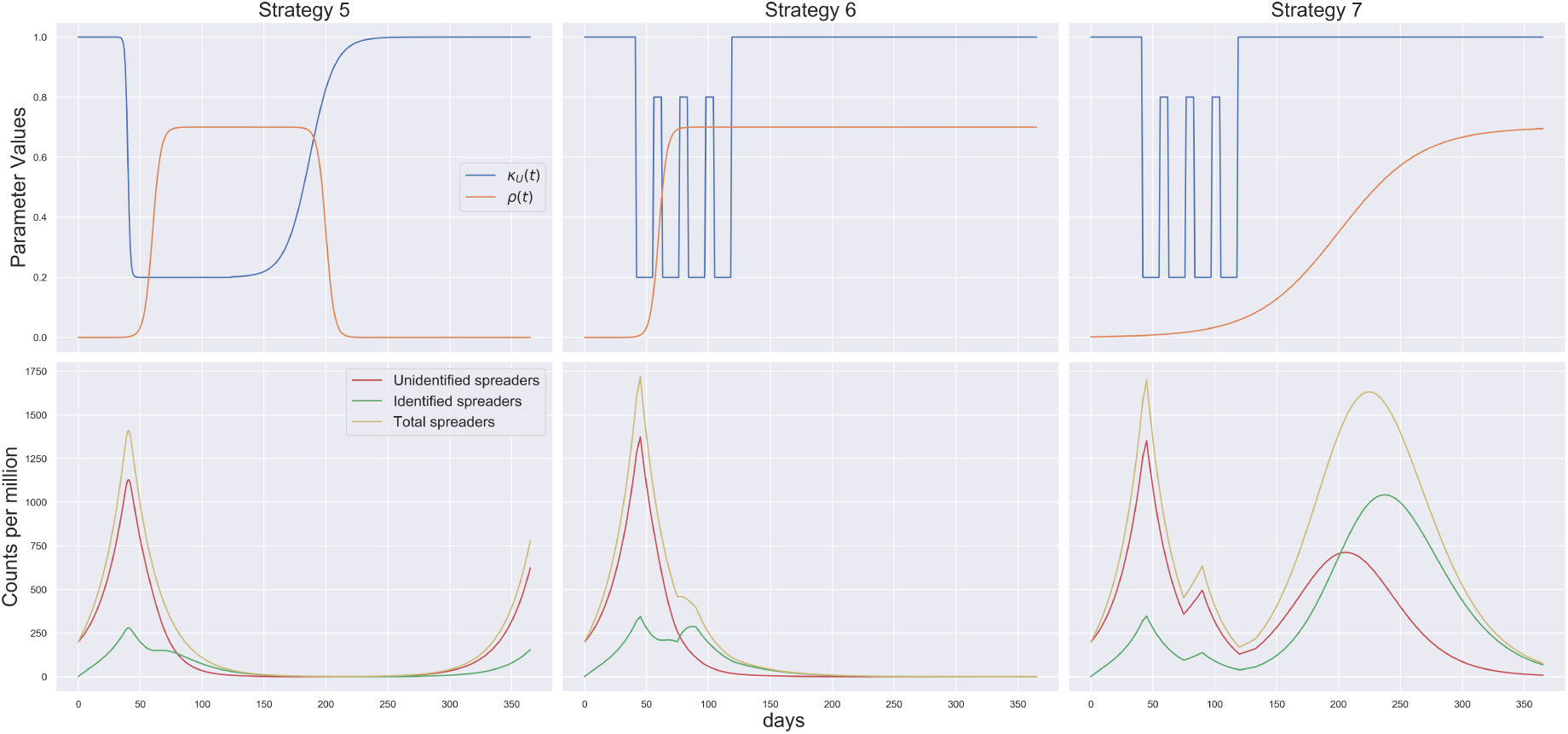
Figure showing parameter values (first row) and number of spreaders (second row) for Strategies 5-7 (represented column-wise)

#### Strategy 6: Periodic lockdown; Early pro-active testing without exit

Periodic lockdown, Early pro-active testing without exit: This is a special case we study motivated by the study of Uri Alon. Here lockdown is implemented on around 40th day. The lockdown is periodic with 1 month of lockdown and 15 days of no lockdown. The lockdown is completely aborted on around 120th day. However, the pro-active testing starts quite early and is continued throughout. This controls the pandemic with no resurgence of cases. (See Figure 7)

#### Strategy 7: Periodic lockdown; Late pro-active testing without exit

Periodic lockdown, late pro-active testing without exit: The lockdown is periodic with 1 month of lockdown and 15 days of no lockdown. The lockdown is completely aborted on around 120th day. Here, the pro active testing starts late, as it is likely to happen in reality. Here there is a resurgence of the pandemic, with more detected cases, since pro-active testing is ramped up. What the plot also shows that without pro active testing in place, the number of unidentified spreaders will be higher for the first wave. As testing is ramped up, more spreaders are identified, so that this peak will be higher. The discussion about what we know and don’t know in terms of spreaders, in relation to the amount of testing done, has played a role in the public debate about the numbers and conclusions drawn. The figure above shows this effect. Comparing with Secario 6 we thus conclude that it is important to ramp up pro-active testing quickly, in order to avoid a second peak. (See Figure 7)

#### Comparing several exit strategies for the pandemic

From Figure 8 we note that four strategies which lack high pro-active testing all have led to a second wave of infections. Also withdrawal of pro active testing at a later time point can also lead to a second wave. This again points out the role of continuing rigorous pro active testing when the lockdown is lifted, until a permanent solution (such as a vaccine) is achieved. (See Figure 7)

**Figure 8:**
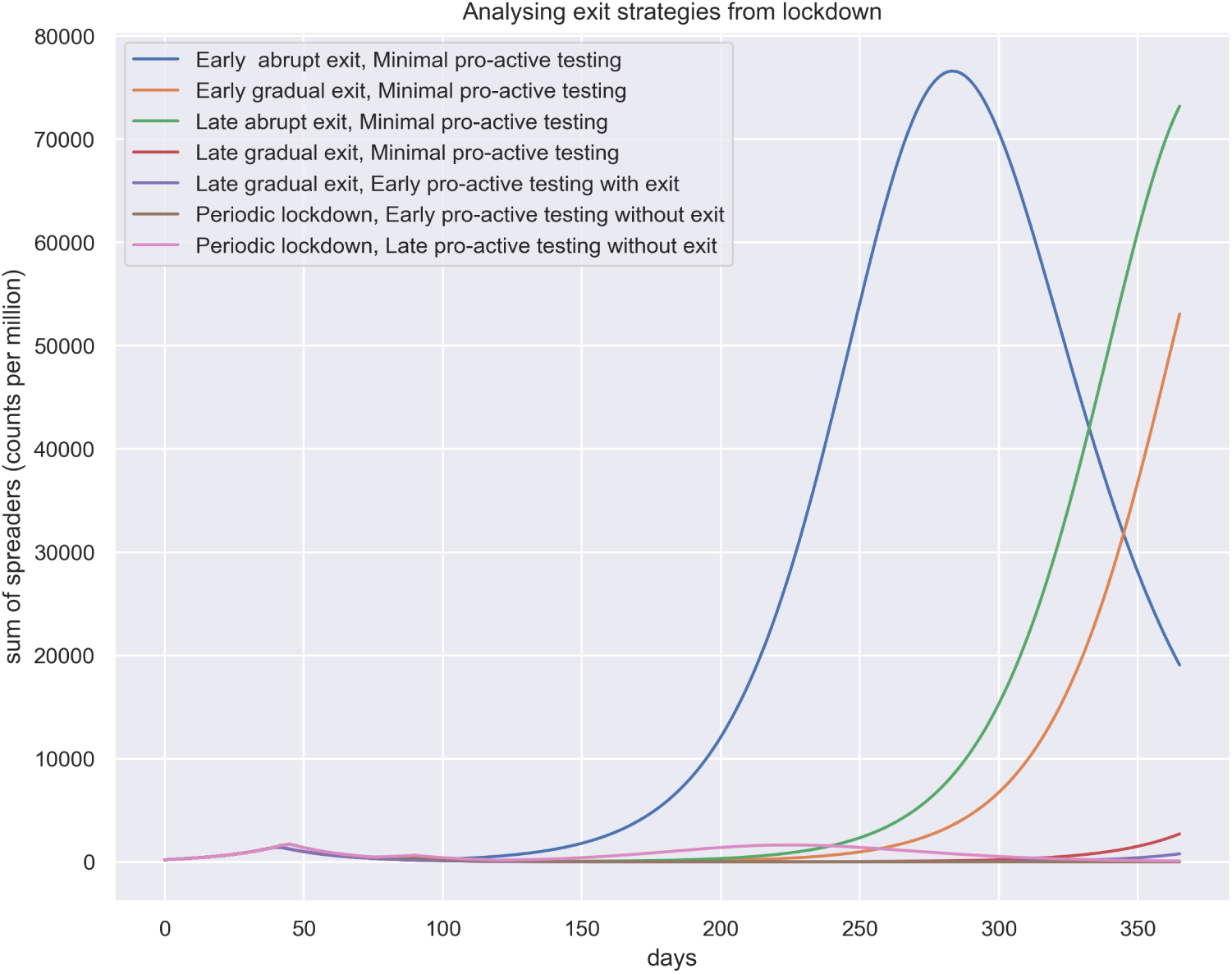
Figure showing for each exit strategy the temporal evolution of spreaders, ie the sum of identified and unidentified spreaders for several strategy.

During the initial phase of the SARS-CoV-2 pandemic, the main question asked with mathematical modelling was about the time and size of the peak of infections.

The peak number of identified spreaders is linked to the number of hospitalisations and ICU beds required. The number of available ICU beds is known and it would be possible to decide upon a threshold that should not be exceeded. The relationship between ICU beds and identified spreaders, can be linked to the case-fatality rate (CFR) - the proportion of diagnosed people who die. Unfortunately, the estimate can vary widely. During the current SARS-CoV-2 pandemic, estimates for the CFR have ranged from 0.1 to 15 percent. The CFR’s denominator - total cases - depends on testing and pro-active testing in particular. The CFR’s numerator - total deaths - depends on the age distribution of a population, the prevalence of preexisting illnesses, and various demographic measures, that vary among countries, states, and cities. This is one example, of how variability and thus uncertainty enters model-based predictions. The reporting of daily new numbers is also known to suffer from various artefacts arising from how the data are collected in different countries.

The second threshold that informs policies is the lower threshold of identified cases, for which relaxation measures can be considered. Here the reproduction number 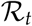 is most widely used. There are numerous approaches to estimate the reproduction number. During the corona crisis, Germany decided on relaxation measures at the end of April, about six weeks after a lockdown was implemented. The number of new infections in Germany, end of April may thus serve as a guide for a lower threshold that should be reached for new identified spreaders, to consider a relaxation of contact restriction measures. Again, estimate of this important parameter induce uncertainty in model-based predictions. We have shown the dynamics of 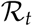 (as calculated in Appendix 5.1) in Figure 9 for some selected case studies. For Policy 1 we oserve expected decrease in Rt if the lockdown is maintained. For Strategy 4 interestingly, while it looks like the cases are low after the first peak, the 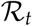 value rises together with an increase in contacts. The 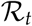 value suggests itself here as a measure for being alerted - a rising 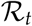 value is an early warning of a second wave. For a periodic lockdown, if pro-active testing is maintained, the 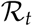 remains within control and thus hinders resurgence of the pandemic.

**Figure 9:**
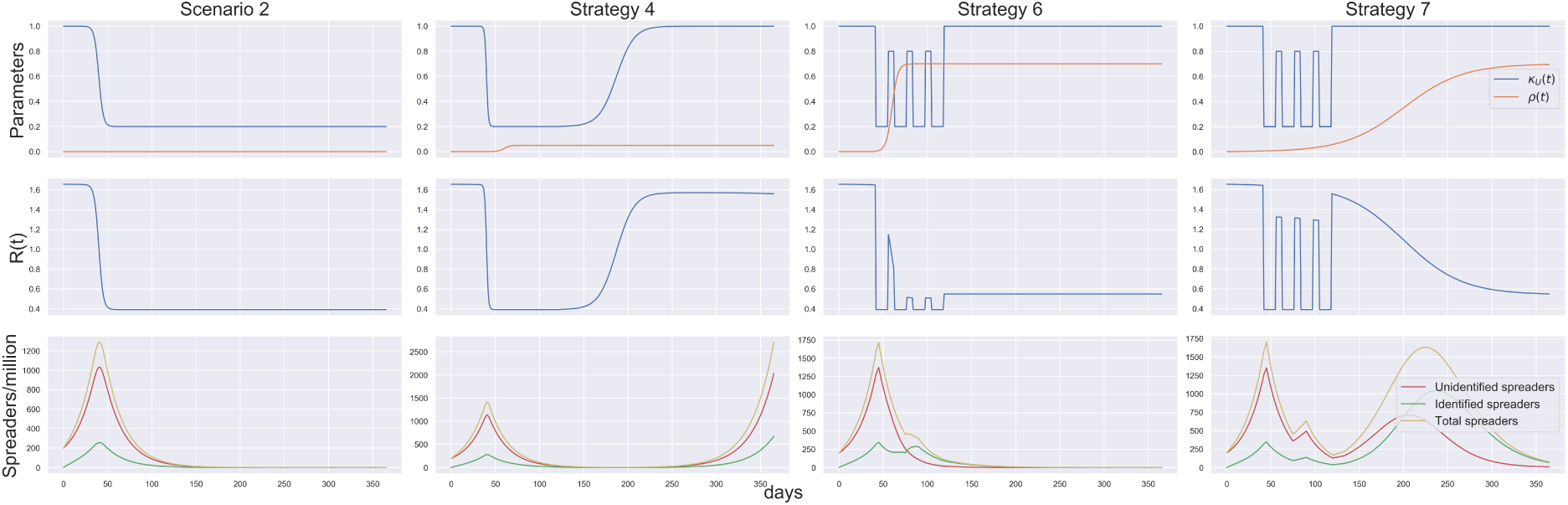
Figure comparing different scenarios, policies and strategies. The top row shows the parameter curves, the middle row shows the dynamics of the reproduction number 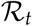 while the bottom row shows the simulated number of spreaders

The use of modeling that we pursued here is one that is not attempting to parameterise the model from actual data. We are therefore not aiming for predictions that are valid for a particular region or country. Such models can still be useful in informing policies and decision making. The focus in then general pattern that emerge from changes to parameters. In this work, we employ this type of modelling, where we do not aim for accurate predictions of case numbers, for a particular region or country. Instead, we focus on parameter changes, linked to testing and contact restriction measures.

More specifically, the SUIR model, presented here, is used to simulate time dependent parameter changes. This allows to investigate strategies that balance pro-active testing with a lockdown. We then focus our discussion on the timing of contract restriction measures and testing. Finally, we study different strategies to exit a lockdown. We show the dynamics of some selected case studies in Figure 9. For our case studies (Scenarios, Policies and Strategies) we quantify the overall number of Identified and Unidentified spreaders by computing the Area Under the Curve (AUC) of the total number of spreaders over our one-year timeline. We then compare percentage changes for several case studies with respect to Scenario 2 (Isolation of identified spreaders; No contact restrictions; No pro-active testing) (See Figure 11) and Policy 1 (Early lockdown; No pro-active testing) (See Figure 12). We choose Scenario 2 for this comparison since, it is the most likely real life scenario considering that no preventive measure is adopted during the pandemic. We choose Scenario 2 for this comparison because, it is the most common entry strategy that has been adopted for the pandemic.

#### Political decision support

Politicians have been looking for criteria that they can use to decide upon a relaxation of contact restrictions. Initially, the 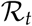 value was brought into the discussion but also created some controversy. Two weeks ago the German government decided on 50 new infections, per week, per 100 000 people, as a criteria. On the 19th of April some states in Germany decided to even lower this threshold (See [Article](https://www.tagesschau.de/inland/coronavirus-deutschland-grenzwert-101.html)). The search for a suitable threshold and single numbered measure to support political decision making is one of the things we would like to investigate further.

If the exit strategy is based on such a threshold, there must be pro-active testing level of around 50 percent to avoid a second peak (in the sense that this threshold is crossed); In other words, there must be testing strategy that 50 percent of all the spreaders are detected and reported. This can prevent the resurgence of a second wave and keep the 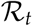 value below the threshold of 1.

## 4 Conclusions

- All our conclusions are based on the assumption that the virus continues to maintain its current potency to spread (*β* remains constant throughout time). Maintaining some degree of contact reduction can make a big difference in case there is a second wave of the pandemic.

In practice this might translate to at least avoiding large gatherings.

- A delayed lockdown has a considerable impact on the number of infections. A delay of only 20 days can increase the number of spreaders at least twofold. In case there is a second wave of the pandemic, early lockdown is thus a very effective measures, provided there is no means of pro-active testing. In our analyses, we assumed an early lockdown to start about 40 days after the first cases.
- If pro-active testing can be employed early, even without any lockdown, the effect of the pandemic can be considerably reduced. This is an important conclusion considering the negative socio-economic effects of a total lockdown.
- Ideally early implementation of lockdown and pro-active testing produces the best results in terms of controlling the number of spreaders. Keeping in mind the possibility of a second wave, we must also note that a late lockdown and late pro-active testing might be much less effective than no lockdown and early pro-active testing. This again emphasizes the importance of developing a pro-active testing strategy before a second wave of the pandemic hits.
- For strategies that are implemented and maintained indefinitely, there is a chance that the pandemic will relapse, within the year after lifting of a short or long lockdown.
- Trends in the reproduction number can be used to forecast the emergence of a second wave. A downturn in the reproduction number does indicate a decline in the number of spreaders. However, since for a value below 1, there are still a considerable number of spreaders, a value below 1 does not give a clear reason to exit a lockdown. A rising curve of the reproduction number, correlates with an increase of contacts, which carries the risk of a second wave. So while the number of identified spreaders may be low, an increase in the reproduction number could serve an alert for a second wave.
- Lockdown (continuous or periodic) can be withdrawn after feasible measures of pro-active testing have been developed. This would be critical to avoid a second peak.
- Considering the socio-economic impact of a total lockdown, a cyclic pattern of lockdown and release, is a promising strategy. A periodic lockdown, along with a high level of pro-active testing maintained continuously after the lockdown is aborted (until the introduction of a vaccine), is a feasible measure to control the pandemic, while maintaining a socio-economic balance.

**Figure 10:**
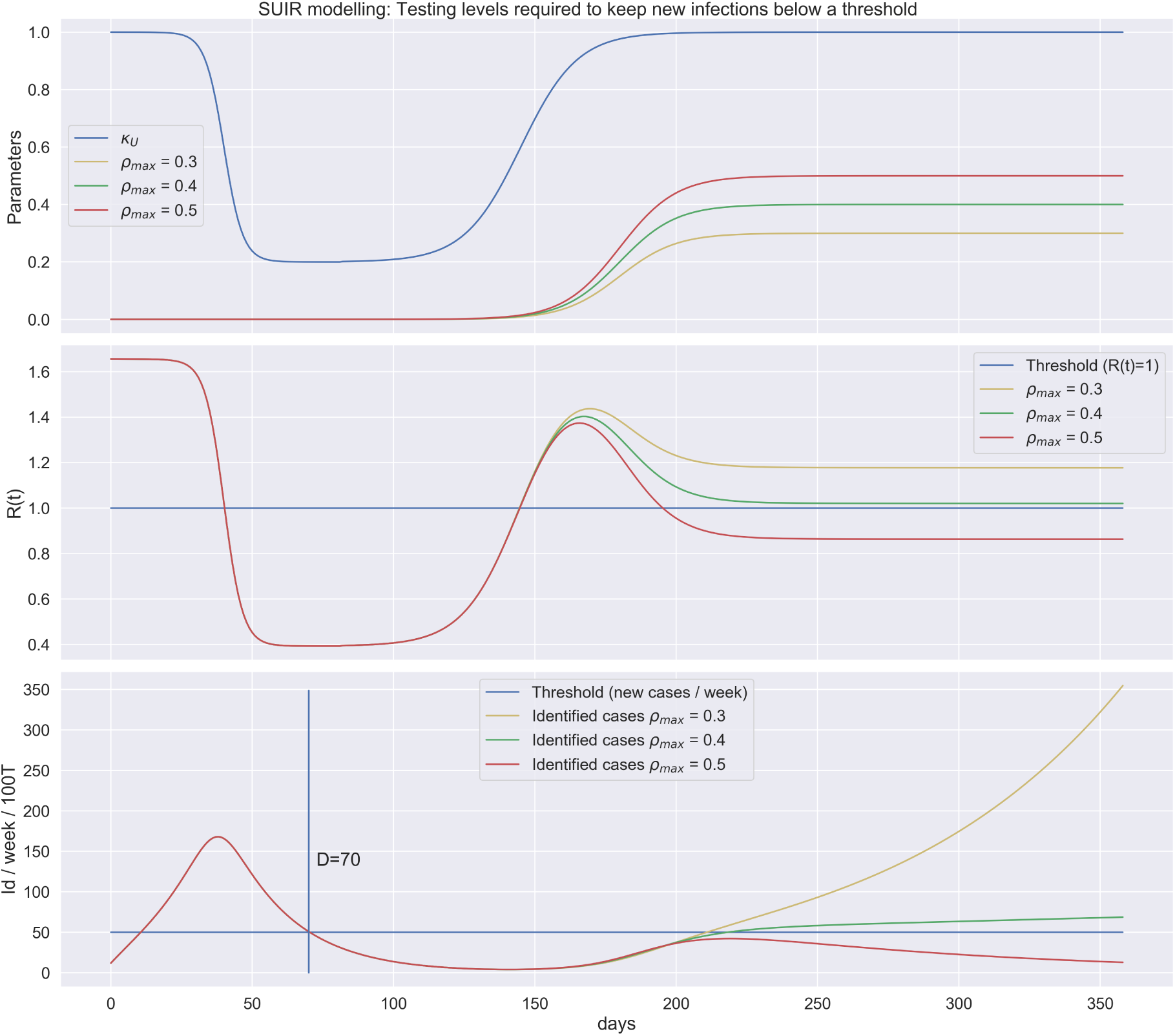
Figure showing dynamics of parameters and 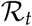 value and number identified cases per 100000 people counted over a 7 day sliding window.

**Figure 11:**
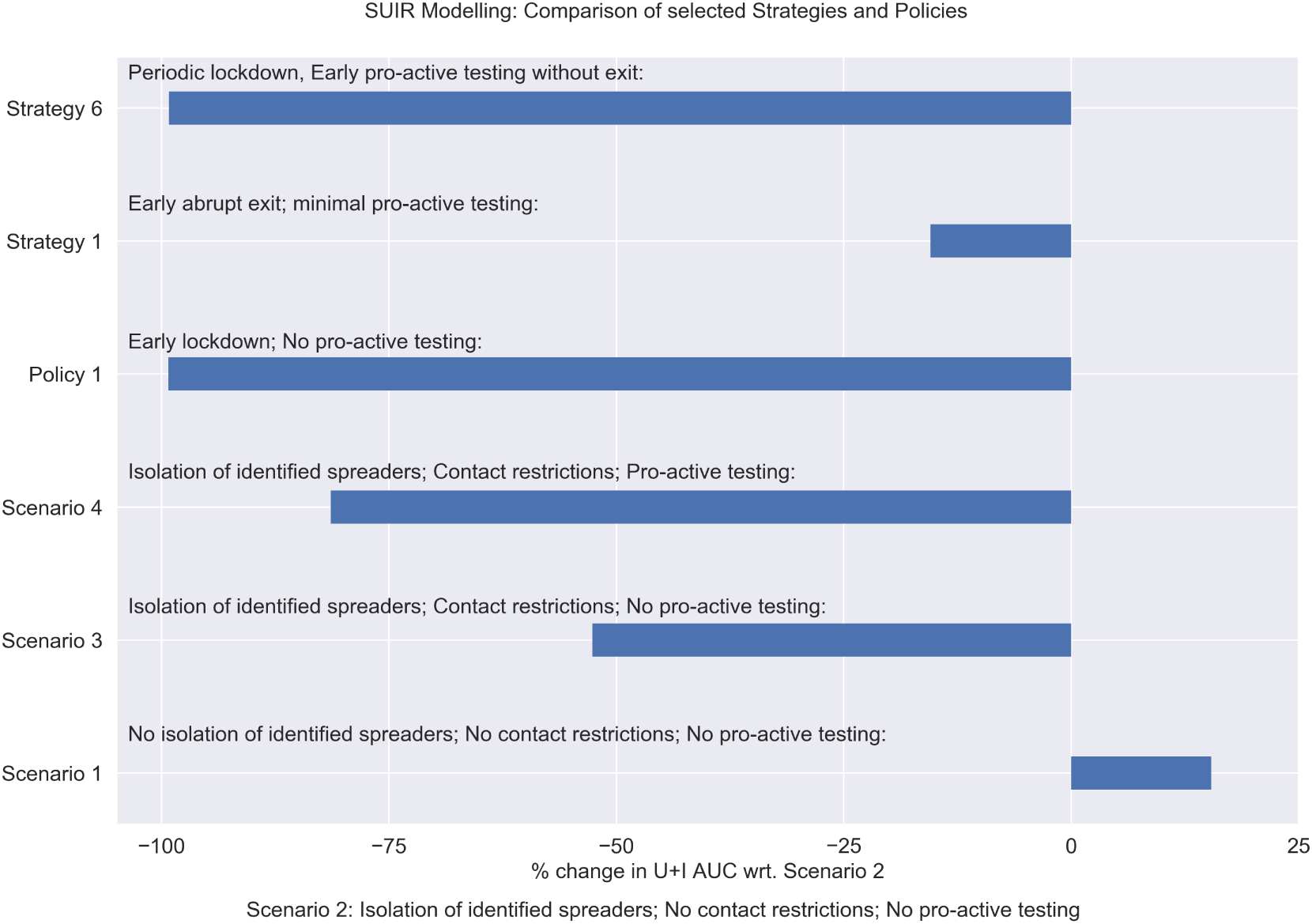
Figure showing percentage change for area under the curve for the total number of spreaders for selected strategies scenarios and policies with respect to Scenario 2

**Figure 12:**
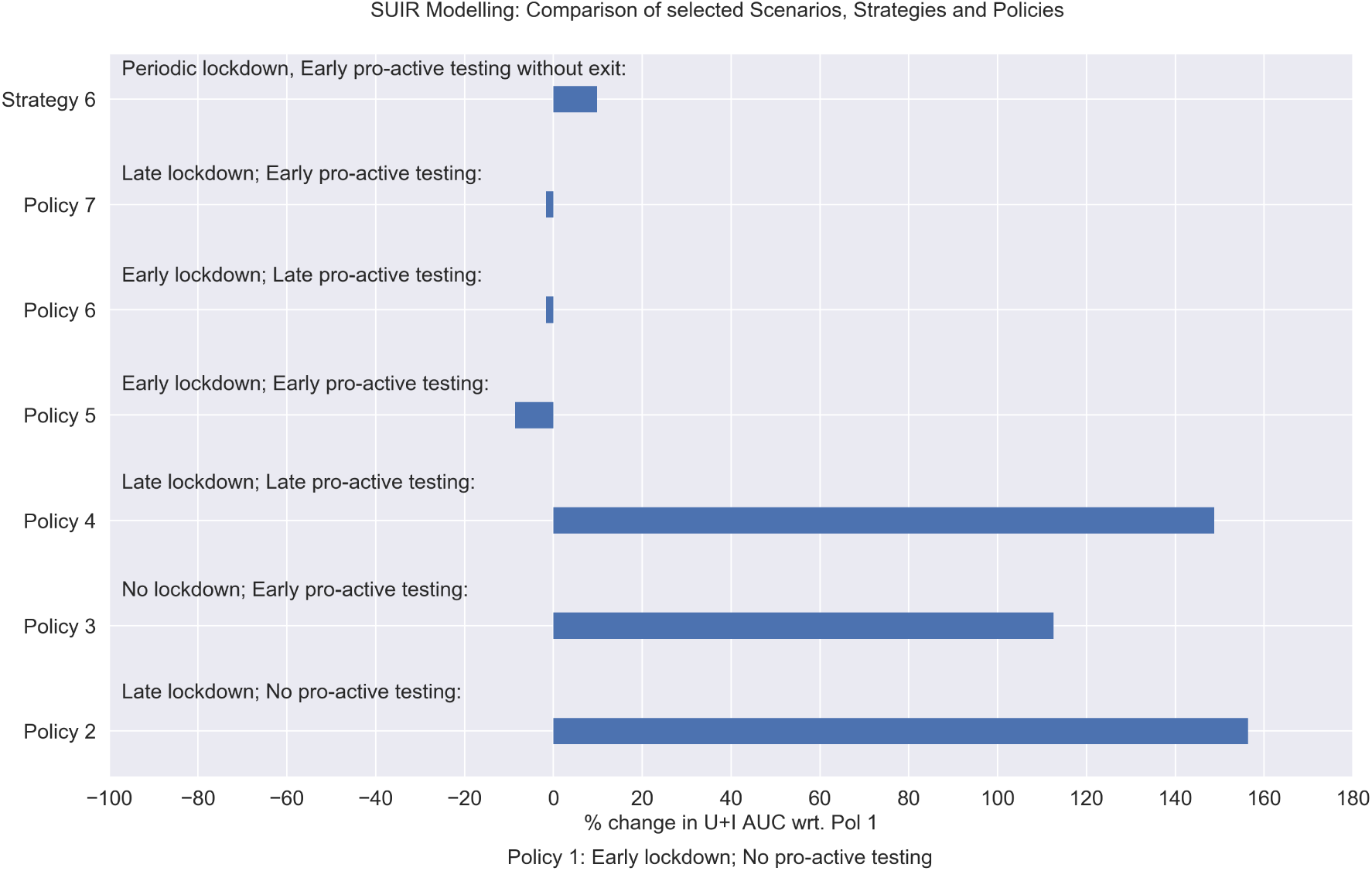
Figure showing percentage change for area under the curve for the total number of spreaders for selected strategies and policies with respect to Policy 1

## Data Availability

The mathematical model and simulation studies presented here, were coded using Python. We used a Jupyter Notebook to to document the derivation of the model in the way it is presented here. The Juypter notebook is available from our Github page at: https://github.com/OlafWolkenhauer/Modelling-the-corona-pandemic. By combining code with text, Jupyter notebooks are making the model and simulation studies transparent and reproducible. Opening the notebook in Google Colab does not even require any installation of Python. Little to no experience in Python is required to use the notebook and explore the SUIR model.

## Materials and methods

The mathematicl model and simulation studies presented here, were coded using Python. We used a Jupyter Notebook to to document the derivation of the model in the way it is presented here. The Juypter notebook is available from our Github page at: Modelling the corona pandemic. By combining code with text, Jupyter notebooks are making the model and simulation studies transparent and reproducible. Opening the notebook in Google Colab does not even require any installation of Python. Little to no experience in Python is required to use the notebook and explore the SUIR model.

## Acknowledgements

We acknowledge valuable discussions in the Working Group Epidemiological Modelling MV (AGEMV) with contributions by Gabriele Doblhammer, Achim Dörre, Aenne Glas, Dagmar Waltemath, Lars Kaderali, and Frank Weber. S.B. was supported by a grant of the Federal German Ministry for Education and Research (BMBF), GB-X MAP (FK 01ZX1709). O.W. acknowledges support by the German Network for Bioinformatics de.NBI & de.STAIR (BMBF FK 031L0106C).

### 5 Appendix

#### 5.1 Calculating reproduction number for SUIR model

The basic reproduction number 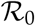 is defined as the expected number of secondary cases produced by a single infection in a completely susceptible population during the initial phase of the pandemic [4]. We follow the next generation matrix method for calculating the 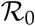 value of our SUIR model [6]. For our SEIR model, the Disease Free Equilibrium (DFE) is given by (*S_eq_*, *E_eq_*, *I_eq_, R_eq_*) = (1,0,0,0). A sub-model that only considers the ‘disease’ compartments, a subset of the equations in the SUIR model, in this case is formed by the subset of Equations 10:

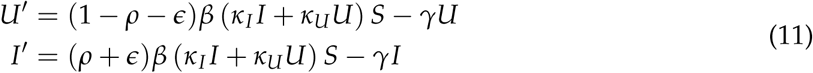

The Equations 11 can be written in the form:

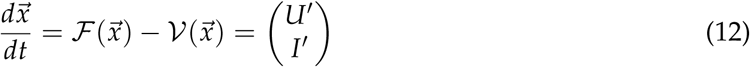

where 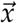 is a vector of the *j* disease compartments; in the SUIR model *j =* 2 because the disease compartments are *U* and *I*. The right hand sides of Equations 11 are therefore contained in the vectors 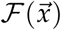 and 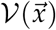 [6]. In our case, 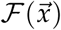 contains any terms that directly lead to new infections entering each compartment *j* and 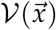 contains any terms that directly lead to infectious individuals leaving each compartment *j* and moving to the compartment 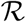. Then, for our model,

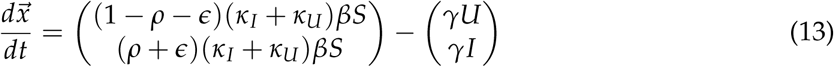

Next, to linearize around the DFE, we calculate the Jacobian matrix for our subset of Equations 11 evaluated at the DFE. Recalling that the DFE is given by (*S_eq_, E_eq_, I_eq_*, *R_eq_*) *=* (1,0,0,0), the Jacobian matrix of the sub-model evaluated at the DFE is given by:

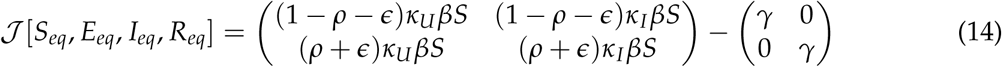

From Equations 13 14 we can write

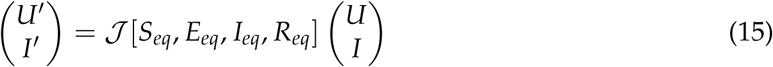

We can rewrite Equation 15 as:

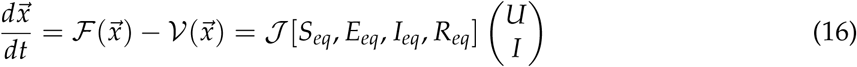

Finally from Equation 16 and 14, we can calculate the value of the next generation matrix 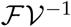

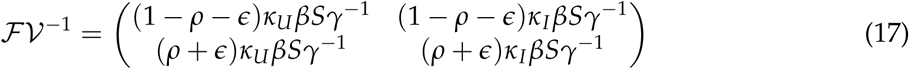

Finally, solving for the highest eigenvalue of the next generation matrix 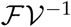 we obtain

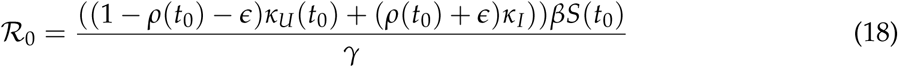

At initial time point *t*_0_, for our model *S* = 1; further assuming κ*_U_* = 1 and *κ_I_* = 1, we note that Equation 18 boils down to 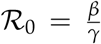 which agrees with the standard SIR model. The basic reproduction number 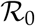 can be generalized over time to obtain time dependent reproduction number 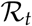 that represents can be interpreted as the expected number of secondary cases produced by a single infection in a completely susceptible population at some time point t during the pandemic [4]. We can further extend Equation 18, to compute 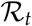:

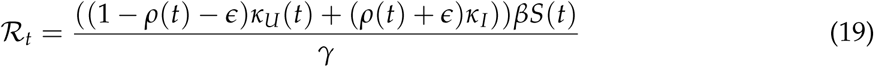

## Notes

### Competing Interest Statement

The authors have declared no competing interest.

